# Viral discovery using untargeted metagenomics and complementary proteomics in plasma cell–rich virus-like tubulointerstitial nephritis after kidney transplantation

**DOI:** 10.64898/2026.11.20.26368586

**Authors:** R. du Long, R. Hofstraat, S. Florquin, N. Claessen, M. Deijs, L. van der Hoek, J. Kers

**Affiliations:** Department of Pathology, Amsterdam University Medical Center, University of Amsterdam, Amsterdam, Netherlands; Van ‘t Hoff Institute for Molecular Sciences, University of Amsterdam, Amsterdam, Netherlands; Department of Pathology, University Medical Center Utrecht, Utrecht University, Utrecht, Netherlands; Laboratory of Experimental Virology, Department of Medical Microbiology and Infection Prevention, Amsterdam UMC, Amsterdam, the Netherlands; Department of Pathology, Systems Pathology Research Group, Leiden Transplant Center, Leiden University Medical Center, Leiden University, Leiden, Netherlands; Department of Pathology, Erasmus MC Transplant Institute, Erasmus Medical Center, Rotterdam, Netherlands

**Keywords:** kidney transplantation, tubulointerstitial nephritis, viral metagenomics, VIDISCA-NGS, BK polyomavirus, virus discovery

## Abstract

**Background:** Kidney allograft biopsies may show plasma cell–rich tubulointerstitial inflammation and tubular epithelial changes suggestive of viral infection despite negative routine testing for known viruses. Whether this plasma cell–rich virus-like tubulointerstitial nephritis (PCR-TIN) is associated with an unrecognized viral agent is unknown.

**Methods:** We retrospectively identified 57 kidney allograft biopsies with a plasma cell–rich tubulointerstitial phenotype. Frozen tissue from 34 biopsies and matched urine from 14 cases underwent virus discovery cDNA-amplified fragment length polymorphism next-generation sequencing (VIDISCA-NGS). BK polyomavirus-associated nephropathy (BKPyVAN) tissue served as a positive control for viral detection in renal tissue. Data-independent acquisition parallel accumulation–serial fragmentation liquid chromatography–tandem mass spectrometry was used to evaluate the feasibility of detecting BK polyomavirus peptides. In an exploratory secondary analysis, host-protein abundance and Hallmark pathway enrichment were compared among 11 PCR-TIN, 91 other rejection, and 20 BKPyVAN biopsies.

**Results:** In positive-control experiments, VIDISCA-NGS detected BK polyomavirus at 13 of 216,879 reads (0.006%) and, after DNase treatment, at 5 of 81,614 reads (0.006%). Across the 34 PCR-TIN biopsies, 3,751,011 reads were generated (mean, 110,324 per biopsy). Sporadic low-level anelloviruses and Merkel cell polyomavirus reads (<10 reads per positive specimen) were detected in five and four biopsies, respectively, but no consistent candidate pathogenic viral agent was identified. Similarly, no candidate pathogenic viral agent associated with PCR-TIN was identified among 409,563 reads from the 14 matched urine samples (mean, 29,255 per sample). Proteomic interrogation did not detect BK polyomavirus peptides even in proven BKPyVAN, rendering this approach technically inconclusive for viral detection in PCR-TIN. Six proteins were significantly more abundant in PCR-TIN than in other rejection, whereas none differed significantly between PCR-TIN and BKPyVAN. Compared with both groups, PCR-TIN showed enrichment of epithelial–mesenchymal transition, interferon-response, allograft-rejection, inflammatory, and unfolded-protein-response programs, whereas oxidative phosphorylation and several metabolic pathways were enriched in the comparator groups.

**Conclusion:** Repeated VIDISCA-NGS of kidney tissue and matched urine did not identify a candidate pathogenic viral agent associated with PCR-TIN. These findings make a prevalent virus detectable by this workflow less likely but do not exclude low-abundance, focal, transient, or methodologically inaccessible infection. Exploratory host proteomics identified inflammatory, cellular-stress, and tissue-remodeling programs associated with PCR-TIN, but these findings characterize the accompanying host response, not its cause.

## Introduction

Tubulointerstitial inflammation is a common pattern of injury in kidney allografts and has a broad differential diagnosis. It may reflect alloimmune injury, infection, drug toxicity, obstruction, ischemic injury, or a combination of processes. Establishing the underlying cause is essential because similar histological patterns may lead to markedly different treatment decisions. In particular, distinguishing viral nephropathy from rejection can be difficult when inflammation and tubular injury dominate the biopsy appearance.

BK polyomavirus (BKPyV) is the best-characterized viral cause of tubulointerstitial nephritis after kidney transplantation. Viral replication in tubular epithelial cells can produce epithelial enlargement, nuclear atypia, cell detachment, tubulitis, and interstitial inflammation, followed by progressive fibrosis and graft dysfunction.^1–4^ Immunohistochemistry for simian virus 40 large T antigen (SV40) and quantitative polymerase chain reaction testing for BKPyV in plasma or urine are therefore routinely used when BKPyV-associated nephropathy (BKPyVAN) is suspected. Other viral infections, including cytomegalovirus (CMV) and varicella-zoster virus (VZV), may also cause clinically relevant disease in transplant recipients.^5,6^ Nevertheless, currently available diagnostic tests identify only *known* pathogens and therefore require a suspected viral target.

A subset of kidney allograft biopsies shows morphological features suggestive of viral injury despite negative routine testing for known nephrotropic viruses. These biopsies may contain dilated tubules with epithelial cell detachment or sloughing, interstitial edema, cytopathic-appearing tubular nuclei, and a prominent plasma cell–rich inflammatory infiltrate. In this study, we refer to this descriptive pattern as plasma cell–rich virus-like tubulointerstitial nephritis (PCR-TIN). The term describes the morphology and the diagnostic question it raises and does not presume either a viral cause or a distinct disease entity. Its appearance differs from the morphological description referred to as plasma cell–rich rejection (PCRR), which is often characterized by a concomitant micro- and macrovascular inflammation without myxoid edema and cytopathic changes.^7^

PCR-TIN poses a clinically important dilemma because viral infection and rejection can overlap morphologically but may require opposing changes in immunosuppression. Plasma cell–rich inflammation and tubulitis may occur in T cell-mediated rejection (TCMR), antibody-mediated rejection (AMR), or mixed rejection.^8^ Conversely, viral infection can induce substantial interstitial inflammation and may coexist with alloimmune injury. Increasing immunosuppression in an unrecognized infection may facilitate viral replication and aggravate tissue injury, whereas reducing immunosuppression in rejection may accelerate alloimmune damage and graft failure.^9–11^ Neither the intensity nor the composition of the inflammatory infiltrate therefore establishes the underlying etiology.

The absence of a known virus on routine testing leaves several possibilities. The biopsy may represent an unusual manifestation of rejection or another non-viral injury process, a known virus may be present below the sensitivity of routine assays, or an as-yet unidentified virus may be involved. The latter possibility is biologically plausible in immunosuppressed recipients, in whom impaired antiviral immunity can permit reactivation or pathogenicity of (opportunistic) viruses that cause little or no disease in immunocompetent hosts. Discovery of an associated virus would have immediate implications for diagnostic testing, epidemiological studies, and the balance between antiviral treatment and immunosuppression. Conversely, a carefully characterized negative discovery study could prevent virus-like morphology from being regarded as sufficient evidence of an occult infection.

Unbiased viral metagenomics provides a means of investigating this question without prespecifying a candidate pathogen. Metagenomic approaches have enabled the identification of previously unrecognized viruses, including a parvovirus associated with chronic tubulointerstitial nephropathy in mice. Studies of the urinary virome have also demonstrated diverse viral populations in kidney transplant recipients and changes associated with immunosuppression.^12^ However, virus discovery in renal tissue is technically challenging because viral nucleic acids may constitute only a minute fraction of the total nucleic-acid content, infection may be spatially focal, and archived specimens may contain degraded genomic material.

Virus discovery cDNA-amplified fragment length polymorphism next-generation sequencing (VIDISCA-NGS) enriches viral sequences through restriction enzyme-mediated fragmentation and amplification of double-stranded (c)DNA. The approach requires less sequencing depth than conventional untargeted metagenomic workflows and has been used to identify a range of previously unrecognized viruses.^13–16^ Examination of kidney tissue allows direct analysis of the affected compartment, while matched urine may contain viral material released from infected tubular epithelial cells and provides a complementary sample from the downstream urinary tract. Protein-based detection could, in principle, offer an orthogonal approach that is less dependent on viral nucleic-acid recovery or sequence amplification, although its feasibility in renal biopsy tissue is uncertain.

We hypothesized that PCR-TIN after kidney transplantation could be associated with an unidentified viral agent. We therefore applied repeated VIDISCA-NGS to kidney allograft tissue and matched urine and used BKPyV-infected kidney tissue to establish whether viral sequences could be recovered from the relevant organ. We additionally evaluated whether high-sensitivity tissue proteomics could detect viral proteins in renal tissue. Our objective was to determine what evidence these complementary approaches provide for or against an unidentified viral etiology of PCR-TIN, while explicitly accounting for the limitations of pathogen non-detection.

## Methods

### Study Design and Case Selection

This retrospective study used kidney allograft biopsies from the Amsterdam UMC of the DEEPGRAFT cohort.^17^ The database comprised 1,055 biopsies obtained between January 1, 2009, and February 1, 2022. All biopsies were initially screened using hematoxylin and eosin (H&E)-stained sections by experienced renal pathologists. Biopsies were eligible for inclusion if they displayed morphological features consistent with virus-like tubulointerstitial nephritis (TIN), defined by dilated tubules with sloughed tubular epithelial cells (TECs), interstitial edema, and mononuclear interstitial infiltrates, frequently with a plasma cell–rich component. Reassessment identified 57 biopsies with a plasma cell–rich tubulointerstitial phenotype, defined by tubular dilation with epithelial cell detachment or sloughing, interstitial edema, and mononuclear interstitial inflammation with a plasma cell–rich component. Biopsies were excluded for downstream analyses if insufficient tissue remained in the formalin-fixed, paraffin-embedded (FFPE) block for molecular analyses, or if an alternative non-viral etiology fully explained the histopathological findings. Routine diagnostic evaluation included SV40 immunohistochemistry for BKPyV. Frozen tissue suitable for VIDISCA-NGS was available from 34 biopsies; matched urine of sufficient quality and quantity was available from 14 of these cases.

Five renal allograft biopsies with suspected or previously demonstrated viral infection were included as controls for viral discovery. These comprised one biopsy with BKPyV infection confirmed by positive SV40 immunohistochemistry; one biopsy with a BKPyV signal detected by NanoString despite negative SV40 immunohistochemistry; and one biopsy each with suspected hantavirus, Merkel cell polyomavirus, or parvovirus B19 infection. Merkel cell polyomavirus infection was suspected based on weak virus-specific immunohistochemical staining, whereas hantavirus and parvovirus B19 were detected by polymerase chain reaction (PCR) performed on FFPE tissue. Peripheral blood samples were not sequenced, as urinary tract–tropic viruses are considered the most likely route of entry to the kidney in the context of viral nephropathy, and the primary aim of the study was to identify viral agents present within renal tissue and the urinary compartment. The study was conducted in accordance with the Declaration of Helsinki and approved by the local ethics and privacy committee (Amsterdam 19.260). Consent was waived because of the retrospective and pseudonymized nature of the data.

### VIDISCA-NGS

Biopsy tissue was crushed in 150 µL of Universal Transport Medium (Copan Diagnostics, Murrieta, California, USA) on ice with disposable pestles (Radnor, Pennsylvania, USA). Cell debris was removed by centrifugation for 10 minutes at 5000 x *g*. Urine samples were processed similarly but without mechanical disruption. Cell-free host DNA was subsequently degraded with 20 U TURBO^TM^ DNase (Thermo Fisher Scientific, Waltham, Massachusetts USA) as described by de Vries et al.^18^ Nucleic acids from viruses were isolated using the Boom method ^19^, and VIDISCA-NGS was performed as described by de Vries et al. with previously described adaptations.^18,20^ Briefly, all nucleic acids were copied into double-stranded DNA via reverse transcription with Superscript II (Thermo Fisher Scientific, Waltham, Massachusetts, USA), non-ribosomal random hexamers and Klenow Fragment (3′→ 5′ exo-) DNA polymerase (New England Biolabs, Ipswich, Massachusetts, USA), and RNase H (Thermo Fisher Scientific, Waltham, Massachusetts, USA).^21^ Double-stranded DNA was fragmented with MseI (T^TAA; New England Biolabs, Ipswich, Massachusetts, USA). MseI-fragmentation in VIDISCA has the advantage that a sample cannot be “over-digested” as fragmentation relies entirely on the presence of restriction enzyme recognition sites and not on the duration of fragmentation. In the subsequent step, adaptors were ligated using T4 DNA ligase (5 U/µL; Thermo Fisher Scientific), while MseI remained active. Adaptor ligation changed the recognition sequence from TTAA to TTAT, thereby preventing repeated cleavage. Size selection with AMPure XP beads (Beckman Coulter, Brea, California, USA) was subsequently performed to remove small DNA fragments prior to a 28-cycle PCR using adaptor-annealing primers. After PCR, the small and large fragments were removed with AMPure XP beads (Beckman Coulter, Brea, California, USA) to select DNA fragments with a length ranging between 200 and 600 bp. DNA was quantified with the Qubit dsDNA HS Assay Kit (Thermo Fisher Scientific, Waltham, Massachusetts, USA) and mixed at equimolar concentrations (56 samples per pool). A bioanalyzer (High Sensitivity DNA Analysis kit, Agilent Genomics, Santa Clara, CA, USA) was used to determine the average nucleotide length of the library pool. Fifty pM DNA was subsequently clonally amplified on beads using the Ion Chef System (Thermo Fisher Scientific, Waltham, Massachusetts, USA). Sequencing was performed on an Ion GeneStudio™ S5 System (Thermo Fisher Scientific, Waltham, Massachusetts, USA) using the Ion 510™ Chip (Thermo Fisher Scientific, Waltham, Massachusetts, USA). Biopsy specimens were subjected to five VIDISCA-NGS runs, of which four yielded usable sequencing data. The four usable runs comprised three standard runs targeting both RNA and DNA viruses and one DNA-focused run in which reverse transcriptase was omitted during the first incubation. Urine samples were subjected to four VIDISCA-NGS runs, of which three yielded usable sequencing data. The three usable urine runs comprised two standard runs and one additional run in which sheep serum was used as a carrier to reduce the inhibitory effect of Boom isolation.^22^ Reads from the four usable biopsy runs and three usable urine runs were pooled per specimen, and all usable runs were included in the final viral-read classification. The viral-control biopsies were analyzed separately, both without DNase pretreatment and after centrifugation and DNase treatment. A pooled output of more than 10,000 sequence reads per specimen was used as the cutoff for further analysis.^20^ All VIDISCA-NGS reads with a minimum length of 45 nucleotides were translated into protein sequences and aligned to a local database of the National Center for Biotechnology Information (NCBI) eukaryotic viral Identical Protein Groups using UBLAST^23^ and the VIDISCA bioinformatics workflow for identification of probable viral reads and background sequence classification^15^.

### Proteomic Feasibility and Comparative Host-Proteome Analysis

Proteomic data were generated from formalin-fixed, paraffin-embedded kidney transplant biopsies using a previously validated liquid chromatography–tandem mass spectrometry workflow, as described by Hofstraat-Boersma et al.^24^ Briefly, proteins were extracted from deparaffinized tissue sections, digested with trypsin, and analyzed using an Evosep One liquid chromatography system coupled to a timsTOF HT mass spectrometer operating in data-independent parallel accumulation–serial fragmentation (diaPASEF) mode. Raw data were processed using DIA-NN without an experimentally generated spectral library. Instead, the UniProt human reference proteome without isoforms was imported together with BK polyomavirus sequences. The viral search was restricted to the BK polyomavirus large T antigen and VP2 proteins to evaluate viral-peptide detectability in a positive-control feasibility analysis. Proven BK polyomavirus–associated nephropathy (BKPyVAN) biopsies served as positive controls, and successful detection of viral peptides in these biopsies was considered necessary for interpreting their absence in PCR-TIN. Identified human proteins were used for the subsequent comparative host-proteome analysis. Additional technical details are provided in the **Supplemental Methods**. Proteomic measurements were initially available from 11 PCR-TIN, 94 other rejection and 20 BKPyVAN biopsies. After exclusion of two auxiliary or repeat measurements and one sample that failed quality control, the final comparative analysis included 11 PCR-TIN, 91 other rejection and 20 BKPyVAN biopsies. Protein groups were retained separately for each comparison when detected in at least 50% of samples in both groups. Protein abundances were log2-transformed and analyzed using linear models with empirical Bayes moderation in limma. Comparisons were defined as PCR-TIN minus other rejection and PCR-TIN minus BKPyVAN. Proteins were considered differentially abundant at a Benjamini–Hochberg false discovery rate below 0.05 and an absolute fold change of at least 1.5. For pathway analysis, proteins were ranked by their moderated limma *t* statistic. Gene symbols were mapped to Entrez identifiers, with the entry showing the largest absolute *t* statistic retained when multiple protein groups mapped to the same identifier. Preranked gene set enrichment analysis was performed using the Molecular Signatures Database Hallmark collection (version 7.5.1). Gene sets containing 10–500 mapped genes were tested, and significance was defined as a Benjamini–Hochberg-adjusted *P*-value < 0.05. Positive normalized enrichment scores indicated enrichment in PCR-TIN, whereas negative scores indicated enrichment in the respective comparator group.

## Results

### Identification and Classification of Biopsies With PCR-TIN

Within the DEEPGRAFT cohort 57 biopsies fulfilled histopathological criteria for a plasma cell–rich tubulointerstitial phenotype (PCR-TIN; **Figure 1A**). Of these, 4 biopsies were classified as pure antibody-mediated rejection, 24 as T cell–mediated rejection grade 1, 6 as T cell–mediated rejection grade 2/3, and 19 as mixed rejection, with the remaining 4 biopsies showing acute tubular injury and/or chronic damage not otherwise specified (**Figure 1B**). PCR-TIN biopsies showed tubular dilation with epithelial cell detachment, nuclear cytopathic changes suggestive of viral infection, (myxoid) interstitial edema, and plasma cell–rich mononuclear infiltrates (**Figure 2**).

**Figure 1.**
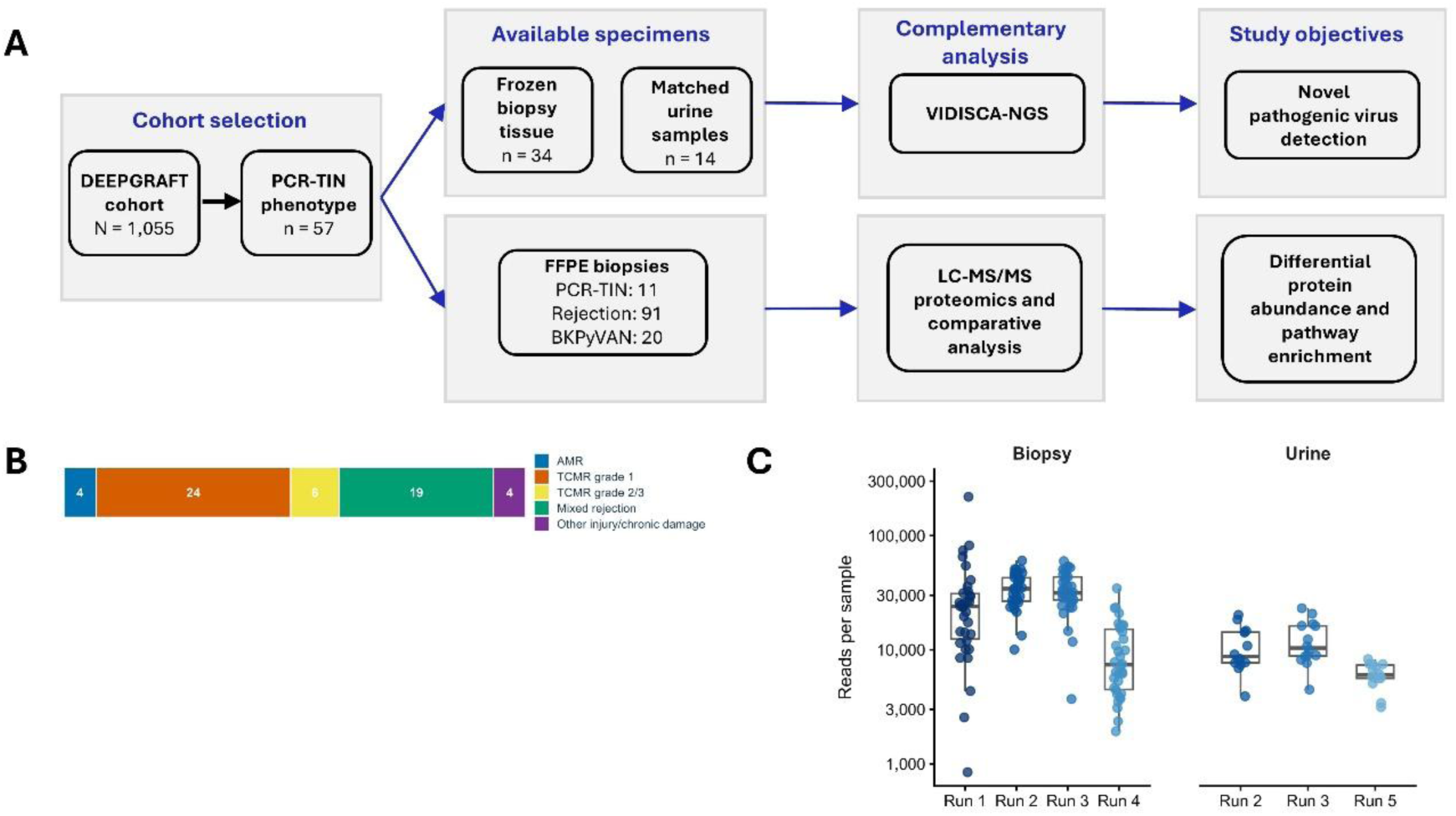
Study Design, Diagnostic Distribution, and VIDISCA-NGS Sequencing Yield in the PCR-TIN Cohort. **A.** From the DEEPGRAFT cohort of 1,055 kidney transplant biopsies, 57 biopsies with a PCR-TIN phenotype were identified. Frozen biopsy tissue was available from 34 biopsies, of which 14 had a matched urine sample. These specimens were analyzed using VIDISCA-NGS to investigate the presence of novel pathogenic viruses. FFPE biopsy proteomic data were available from 11 PCR-TIN, 91 other rejection, and 20 BKPyVAN biopsies. LC–MS/MS proteomics was used to compare protein abundance and Hallmark pathway enrichment between PCR-TIN and the two comparison groups. **B.** Distribution of the diagnostic categories among the 57 biopsies with a PCR-TIN phenotype. Numbers within the bar indicate the number of biopsies in each category. **C.** VIDISCA-NGS reads obtained across four biopsy runs and three urine runs. Points represent individual specimens; boxes indicate the median and interquartile range, and whiskers extend to 1.5 times the interquartile range. Read counts are displayed on a logarithmic scale. *Abbreviations:* AMR, antibody-mediated rejection; BKPyVAN, BK polyomavirus–associated nephropathy; FFPE, formalin-fixed, paraffin-embedded; LC–MS/MS, liquid chromatography–tandem mass spectrometry; PCR-TIN, plasma cell-rich tubulointerstitial nephritis; TCMR, T cell–mediated rejection; VIDISCA-NGS, virus discovery cDNA-amplified fragment-length polymorphism next-generation sequencing.

**Figure 2.**
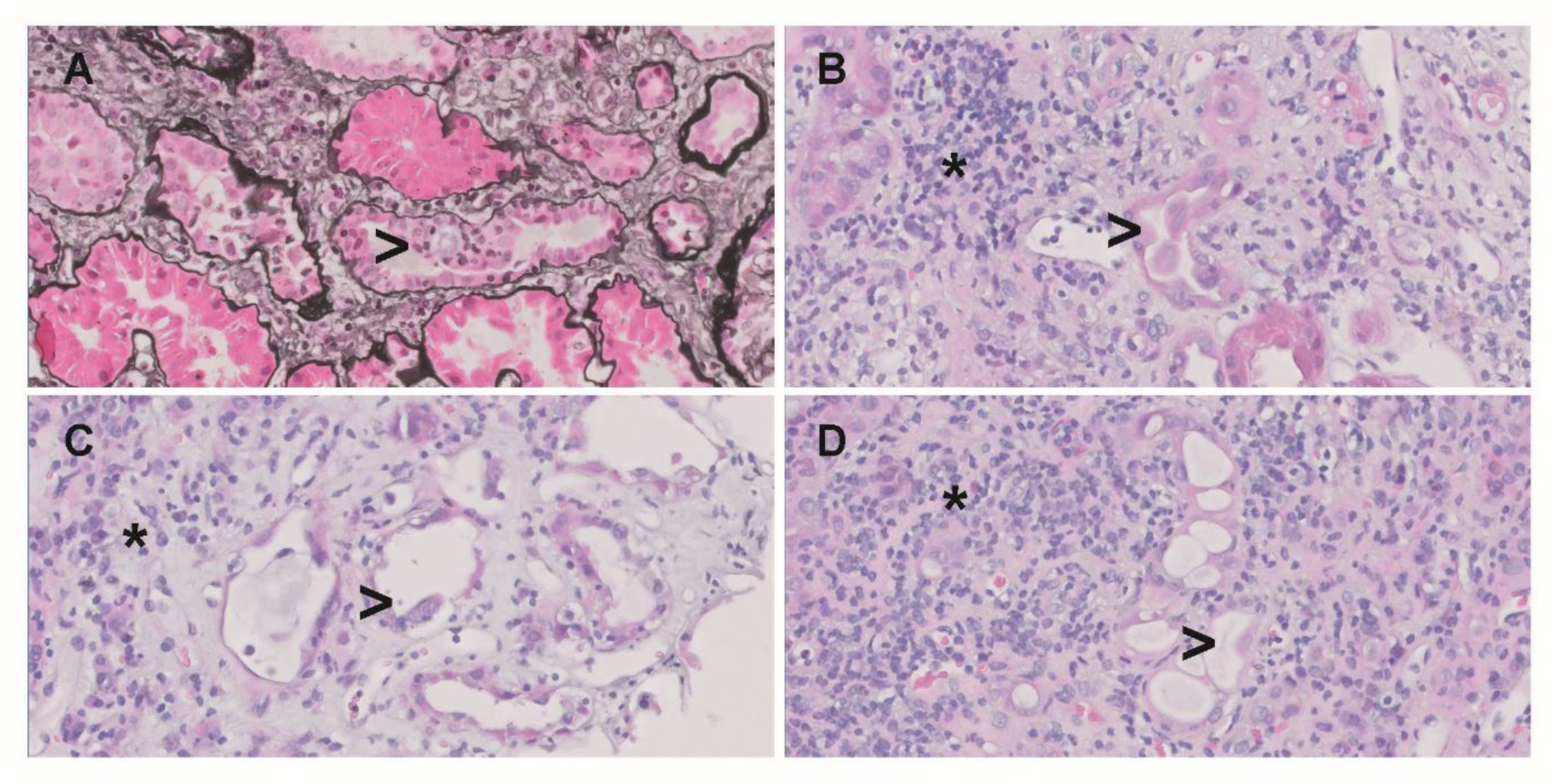
Histopathological Features of Plasma Cell-Rich Tubulointerstitial Nephritis. Representative kidney allograft biopsies showing the characteristic morphological features of PCR-TIN. **A.** Silver-stained section showing a dilated tubule containing sloughed tubular epithelial cells. **B–D.** Hematoxylin and eosin-stained sections demonstrating tubular epithelial injury with tubular dilatation and intraluminal sloughed epithelial cells (arrow heads), accompanied by interstitial edema and plasma cell-rich inflammation (asterisk). *Abbreviations*: PCR-TIN, plasma cell-rich tubulointerstitial nephritis.

### VIDISCA-NGS Detects BKPyV in Positive-Control Kidney Tissue

In the positive-control experiments, BKPyV was detected in one biopsy with proven BKPyVAN, both without DNase treatment (13 BKPyV reads among 216,879 total reads; 0.006%) and after DNase treatment (5 BKPyV reads among 81,614 total reads; 0.006%). The other control samples generated pooled sequencing outputs above the predefined threshold of 10,000 reads, but the expected hantavirus, Merkel cell polyomavirus, and parvovirus B19 sequences were not detected. Thus, successful viral detection in kidney tissue was demonstrated only for BKPyV. BKPyV represented a very small proportion of the total sequencing output and remained detectable after DNase treatment, with an identical proportion of BKPyV reads in both preparations.

### No Candidate Novel Pathogenic Virus is Identified in PCR-TIN Tissue of Urine

The four usable VIDISCA-NGS runs from the 34 PCR-TIN biopsies generated 3,751,011 pooled reads, with a mean of 110,324 reads per biopsy. All biopsy specimens exceeded the predefined pooled-output threshold of 10,000 reads. Low numbers of anelloviruses reads were detected in five biopsies (5/34; 14.7%), and Merkel cell polyomavirus reads were detected in four biopsies (4/34; 11.8%). Each positive finding comprised fewer than 10 viral reads per specimen. Most of these low-level viral detections were obtained in the DNA-focused VIDISCA run, consistent with the DNA genomes of these viruses. The detections were sporadic, and neither virus was detected throughout the cohort. No credible candidate pathogenic viral agent associated with PCR-TIN was identified. The three usable runs from the 14 matched urine samples generated 409,563 pooled reads, with a mean of 29,255 reads per sample (**Figure 1C; Supplementary Table S1**). All urine specimens exceeded the predefined pooled-output threshold. No candidate pathogenic viral agent associated with PCR-TIN was identified in urine.

### Proteomics Does not Detect BKPyV in the Positive Controls

Targeted interrogation of the diaPASEF data did not detect peptides derived from BKPyV large T antigen or VP2 in either PCR-TIN or proven BKPyVAN biopsies. Because viral peptides were not recovered from the positive-control BKPyVAN samples, the sensitivity of this approach for detecting BKPyV proteins in archival FFPE kidney tissue could not be established. The absence of these viral peptides in PCR-TIN was therefore considered technically inconclusive and was not interpreted as evidence against a viral cause.

### Exploratory Comparative Host-Proteome Analysis

Comparative host-proteome analysis included PCR-TIN (n = 11), other rejection (n = 91), and BKPyVAN (n = 20) biopsies. Six proteins (TXNDC5, PRDX4, EMILIN2, MZB1, FKBP11, and POSTN) were significantly more abundant in PCR-TIN than in other rejection, whereas no individual proteins differed significantly between PCR-TIN and BKPyVAN after correction for multiple testing (**Supplementary Figure S1** and **Supplementary Tables S2 and S3**). MZB1 showed the largest difference, with an approximately 8.2-fold higher abundance in PCR-TIN.

Hallmark gene set enrichment analysis identified 23 significantly enriched pathways in the comparison with other rejection and 19 in the comparison with BKPyVAN (**Supplementary Figure S2 and Supplementary Tables S4 and S5**). Fifteen pathways were significant in both comparisons. PCR-TIN showed consistent enrichment of epithelial–mesenchymal transition, interferon-γ and interferon-α responses, allograft rejection, inflammatory response, IL-6–JAK–STAT3 signaling, unfolded protein response, G2/M checkpoint, and MYC targets V1. Conversely, oxidative phosphorylation, fatty acid metabolism, xenobiotic metabolism, adipogenesis, bile acid metabolism, and peroxisomal pathways were consistently enriched in both other rejection and BKPyVAN relative to PCR-TIN. Additional comparison-specific findings included enrichment of TNF-α signaling via NF-κB, complement, and angiogenesis in PCR-TIN versus BKPyVAN.

## Discussion

In this study, repeated untargeted viral metagenomic sequencing, including additional DNA-focused biopsy sequencing and carrier-assisted urine sequencing, identified sporadic low-level anelloviruses and Merkel cell polyomavirus reads but did not identify a consistent candidate pathogenic viral agent associated with PCR-TIN in 34 kidney allograft biopsies or 14 matched urine samples. VIDISCA-NGS recovered BKPyV from positive-control kidney tissue, demonstrating that viral sequences could be detected in the affected organ. However, the low proportion of BKPyV reads illustrates the challenging signal-to-background ratio in solid tissue. Our findings therefore provide no evidence for a prevalent, sufficiently abundant viral agent accessible to this workflow, but they do not establish that PCR-TIN has no viral cause.

The low-level viral detections did not provide evidence for an etiological association with PCR-TIN. Anelloviruses reads were found in five biopsies and Merkel cell polyomavirus reads in four, with fewer than 10 reads in each positive specimen. Neither virus was detected in urine, and no viral agent was consistently present across the cohort. Anelloviruses are common components of the human virome, while Merkel cell polyomavirus is frequently present on human skin and may be introduced during tissue sampling or processing. These sporadic, very low-abundance findings were therefore not considered evidence of a pathogenic virus underlying the PCR-TIN phenotype. Most of the additional viral detections were obtained through the DNA-focused VIDISCA run. This demonstrates that the modified run contributed information beyond the standard runs, particularly for DNA viruses, but the detection of viral sequences alone does not establish tissue infection or causality.

The negative result is nevertheless clinically informative. Virus-like morphology alone should not be regarded as evidence of occult infection or used to justify pathogen-specific testing or treatment. Conversely, non-detection is not proof of absence. A virus could have been present below the detection limit, confined to an unsampled focus, no longer present at the time of biopsy, restricted to another compartment, or inadequately recovered because of pre-analytical degradation or assay limitations. PCR-TIN may also be etiologically heterogeneous, with a rare viral cause present in only a small subset of biopsies.

The metagenomic analysis was strengthened by direct examination of kidney tissue, pooling of repeated sequencing runs, inclusion of matched urine for a subset of cases, use of modified runs intended to improve recovery of DNA viruses from tissue and nucleic acids from urine, and detection of BKPyV in infected renal tissue. However, although the other virus-control samples generated sequencing outputs above the predefined threshold, the expected hantavirus, Merkel cell polyomavirus, and parvovirus B19 sequences were not recovered. Adequate overall sequencing yield therefore did not ensure recovery of the expected viral sequences, and successful detection was demonstrated only for BKPyV. A positive control for BKPyV does not establish equivalent sensitivity for viruses with different genome structures, abundance levels, or sequence compositions. VIDISCA-NGS depends on nucleic-acid preservation, double-stranded (c)DNA generation, restriction-enzyme recognition sites, fragment recovery, and database similarity. Limitations at any of these stages may impair detection of a low-copy or highly divergent virus. Differences between the tissue used for prior diagnostic testing and the tissue analyzed by VIDISCA-NGS, together with focal viral distribution, may also have contributed to non-detection in the other control biopsies.

The proteomic analysis addressed two separate questions. First, we evaluated whether BKPyV peptides could be recovered from archival kidney-biopsy material. Neither large T antigen nor VP2 was detected in proven BKPyVAN biopsies. Because the method did not meet its positive-control requirement, the absence of these peptides in PCR-TIN was technically inconclusive and cannot be considered additional evidence against a viral cause. Second, exploratory analysis of the human proteome identified differences between PCR-TIN and the comparator groups. Six proteins—TXNDC5, PRDX4, EMILIN2, MZB1, FKBP11, and POSTN—were more abundant in PCR-TIN than in other rejection phenotypes. Several are involved in protein folding and endoplasmic-reticulum homeostasis or extracellular-matrix organization and tissue remodeling. MZB1 showed the largest difference, although its lower detection frequency in rejection indicates that this finding reflects both more frequent detection and higher measured abundance in PCR-TIN. No individual proteins remained significantly different between PCR-TIN and BKPyVAN after correction for multiple testing.

Hallmark analysis demonstrated coordinated pathway-level differences in both comparisons. Relative to other rejection and BKPyVAN, PCR-TIN showed enrichment of epithelial–mesenchymal transition, interferon response, allograft rejection, inflammatory-response, and unfolded-protein-response programs. Conversely, oxidative phosphorylation and several metabolic pathways were enriched in the comparator groups. Together, these findings suggest that PCR-TIN is accompanied by inflammation, cellular stress, and tissue remodeling. However, these pathways are not specific to infectious or alloimmune injury, and epithelial–mesenchymal-transition enrichment in tissue-level proteomic data should be interpreted as a remodeling-associated program rather than evidence of complete phenotypic transition in tubular epithelial cells.

Many PCR-TIN biopsies also fulfilled Banff rejection criteria, suggesting that PCR-TIN may represent a morphological pattern occurring across several clinicopathological diagnoses rather than a distinct disease entity. The observed proteomic differences could reflect variation in inflammatory-cell composition, plasma-cell abundance, injury chronicity, treatment exposure, or other clinical characteristics. The small PCR-TIN group, unequal group sizes, and lack of adjustment for these potential confounders mean that the proteomic findings should be considered exploratory and require independent validation.

Additional limitations include the availability of tissue for only 34 of 57 phenotype cases and urine for 14, creating potential selection based on specimen availability. Peripheral blood was not analyzed, and the study was not designed to estimate a formal upper bound for the prevalence of a rare or heterogeneous viral cause. Moreover, sequencing depth does not translate into a uniform detection probability across viruses and specimens. The inconsistent recovery of the expected viruses from the control biopsies further precluded estimation of virus-independent detection sensitivity for the VIDISCA-NGS workflow. Future studies should prospectively collect fresh-frozen tissue, urine, and blood; incorporate orthogonal nucleic-acid methods; use spatial approaches to target cytopathic-appearing tubules; and include standardized positive controls and spike-in detection limits. Targeted or spatial protein assays may help identify the cells and compartments responsible for the inflammatory, cellular-stress, and remodeling-associated signals observed here.

In conclusion, VIDISCA-NGS did not identify a candidate pathogenic viral agent associated with PCR-TIN in kidney tissue or matched urine. These findings reduce the likelihood that a common, readily detectable virus underlies the phenotype but do not exclude focal, low-level, transient, rare, or methodologically inaccessible infection. Viral-peptide detection by proteomics was technically inconclusive, whereas exploratory host proteomics identified inflammatory, cellular-stress, and tissue-remodeling programs without establishing their cause. The etiology of PCR-TIN therefore remains unresolved, and its virus-like morphology should be interpreted together with validated pathogen testing and the broader clinicopathological context.

## Data Availability

All data produced in the present study are available upon reasonable request to the authors

## Author contributions

Dr. Florquin, Dr. Kers, Dr. Deijs, and Dr. van der Hoek designed the study. Practical experiments were performed at the Department of Pathology and Laboratory of Experimental Virology by Ms. Claessen, Dr. Deijs and Dr. van der Hoek. Ms. du Long performed data collection and processing. Sample preparation, practical experiments and data processing for the proteomics analysis were performed by Ms. Hofstraat. Ms. du Long wrote the original draft of the manuscript. The final version of the manuscript was revised and approved by all authors.

