## Supplemental Material for "Viral discovery using untargeted metagenomics and complementary proteomics in plasma cell–rich virus-like tubulointerstitial nephritis after kidney transplantation"

Du Long et al. 2026

**Supplementary material**

### Supplementary Methods

#### Tissue Proteomics and Data Acquisition

Proteomic data were obtained using the formalin-fixed, paraffin-embedded kidney biopsy workflow developed and technically validated by Hofstraat-Boersma et al.[1] Briefly, tissue sections were collected directly into microcentrifuge tubes and deparaffinized using sequential xylene and ethanol incubations. Following heat-induced protein extraction, proteins were reduced with dithiothreitol, alkylated with iodoacetamide, and digested with trypsin. The resulting peptides were purified by solid-phase extraction, freeze-dried, and stored at −80 °C until instrumental analysis. Peptides were analyzed using an Evosep One liquid chromatography system with the 60-samples-per-day method and a timsTOF HT mass spectrometer operating in diaPASEF mode. The chromatographic method used a 21-minute gradient and a 24-minute total cycle time. Quality-control samples were included at regular intervals during acquisition. Full details of tissue preparation, chromatography, mass-spectrometric settings, and technical validation have been reported previously.[1] Raw DIA-PASEF data were processed using DIA-NN. A sequence database based on the human UniProt reference proteome without isoforms was supplemented with BK polyomavirus protein sequences to permit assessment of viral-peptide detection. Protein quantities used in the comparative host analysis corresponded to DIA-NN gene-group abundances. Retention-time-dependent and label-free quantification normalization were performed during DIA-NN processing as described previously.[1]

#### Sample Selection and Quality Control

The comparative analysis included biopsies classified as PCR-TIN, other rejection, or BKPyVAN. Eleven predefined PCR-TIN biopsies were included. Auxiliary or repeat measurements RA0657_30_TR and RA0969_30_MR were excluded to retain independent measurements. Sample RA0769_TR was excluded because 99.2% of protein-group measurements were missing, leaving only 49 detected protein groups. The final analysis therefore comprised 11 PCR-TIN, 91 other rejection, and 20 BKPyVAN biopsies. Separate protein matrices were created for PCR-TIN versus other rejection and PCR-TIN versus BKPyVAN. Within each comparison, protein groups were retained if they were detected in at least 50% of samples in each group. This required detection in at least 6 of 11 PCR-TIN and 46 of 91 rejection biopsies for the rejection comparison, and in at least 6 of 11 PCR-TIN and 10 of 20 BKPyVAN biopsies for the BKPyVAN comparison. After filtering, 4,285 protein groups were retained for PCR-TIN versus rejection and 4,356 for PCR-TIN versus BKPyVAN. Missing measurements were retained as missing and were not imputed for the comparative analysis.

**Differential Protein-Abundance Analysis**

Protein abundances were log2-transformed before statistical analysis. Differential abundance was assessed separately for PCR-TIN versus other rejection and PCR-TIN versus BKPyVAN using the limma package in R. For each comparison, a no-intercept design matrix was constructed, and the contrast was specified as PCR-TIN minus the relevant comparator. Protein-wise linear models were fitted using lmFit, followed by contrast estimation and empirical Bayes moderation using eBayes with trend and robust estimation enabled.

Multiple testing was controlled using the Benjamini-Hochberg method. Proteins were classified as differentially abundant when the adjusted *P* value was below 0.05 and absolute log2 fold change ≥ log2(1.5), approximately **0.585**. Positive log2 fold changes indicated higher abundance in PCR-TIN. Analyses were performed using protein-intensity data; DESeq2 was not used because these values were continuous, normalized protein abundances rather than sequencing read counts.

**Hallmark Gene Set Enrichment Analysis**

Preranked gene set enrichment analysis was based on moderated *t* statistics from the corresponding limma models. Protein groups containing multiple gene symbols were separated, and suffixes introduced solely to ensure unique protein-group names were removed before annotation. Human gene symbols were mapped to Entrez identifiers using org.Hs.eg.db. When more than one protein-group entry mapped to the same Entrez identifier, the entry with the largest absolute moderated *t* statistic was retained. This produced ranked lists of 4,117 and 4,189 unique Entrez identifiers for the rejection and BKPyVAN comparisons, respectively. Preranked gene set enrichment analysis (GSEA) was performed using clusterProfiler and the Molecular Signatures Database (MSigDB) Hallmark gene-set collection, version 7.5.1. The fgsea implementation was used with a fixed random seed of 123, a minimum gene-set size of 10, a maximum size of 500, and no preliminary nominal *P*-value filtering. Adjusted *P* values were calculated across all tested Hallmark pathways using the Benjamini-Hochberg method. Pathways with adjusted *P* values below 0.05 were considered significant. Because the rankings were derived from contrasts defined as PCR-TIN minus the comparator, positive normalized enrichment scores indicated enrichment in PCR-TIN, whereas negative scores indicated enrichment in other rejection or BKPyVAN.

#### Viral-Peptide Detection

To evaluate whether viral proteins could be detected in archival kidney-biopsy material, diaPASEF data from proven BKPyVAN biopsies were used as positive controls. The sequence database used for DIA-NN processing included BK polyomavirus large T antigen and VP2 proteins in addition to the human reference proteome. Detection of BK polyomavirus-derived peptides in proven BKPyVAN was considered necessary to establish the technical interpretability of negative viral-protein findings in PCR-TIN.

### Supplementary Tables S1 – S5

The five supplementary tables are provided in a separate Excel workbook accompanying this preprint, with one worksheet per table.

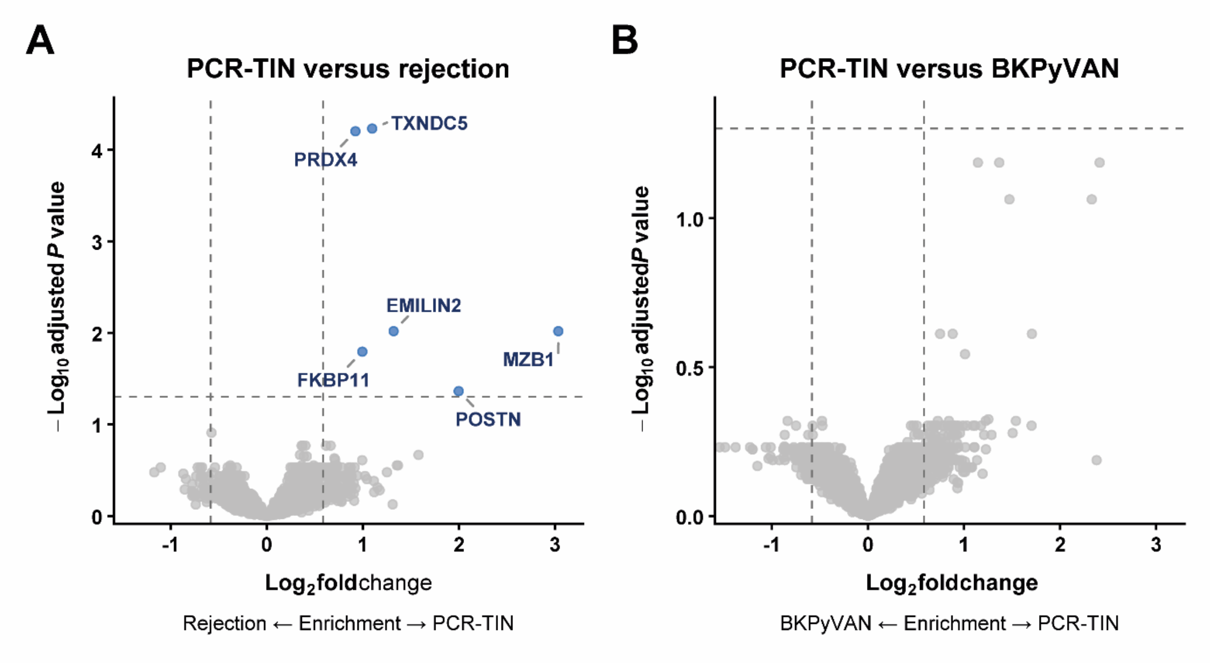


**Supplementary Figure S1. Differential Protein Abundance in PCR-TIN Compared With Rejection and BKPyVAN.** Volcano plots show differential protein abundance **A.** PCR-TIN versus rejection. **B.** PCR-TIN versus BKPyVAN. Each point represents a protein group. Positive $\log_2$ fold changes indicate higher abundance in PCR-TIN, whereas negative values indicate higher abundance in the respective comparison group. Differentially abundant proteins were defined by a Benjamini–Hochberg FDR <0.05 and an absolute fold change ≥1.5 and are shown in teal; nonsignificant proteins are shown in grey. Significant proteins are labelled in panel A. No proteins met the significance criteria in panel B. Vertical dashed lines indicate the fold-change threshold, and horizontal dashed lines indicate an FDR of 0.05. Panels have independent y-axis scales.

**Abbreviations**: BKPyVAN, BK polyomavirus–associated nephropathy; FDR, false discovery rate; PCR-TIN, plasma cell–rich virus-like tubulointerstitial nephritis.

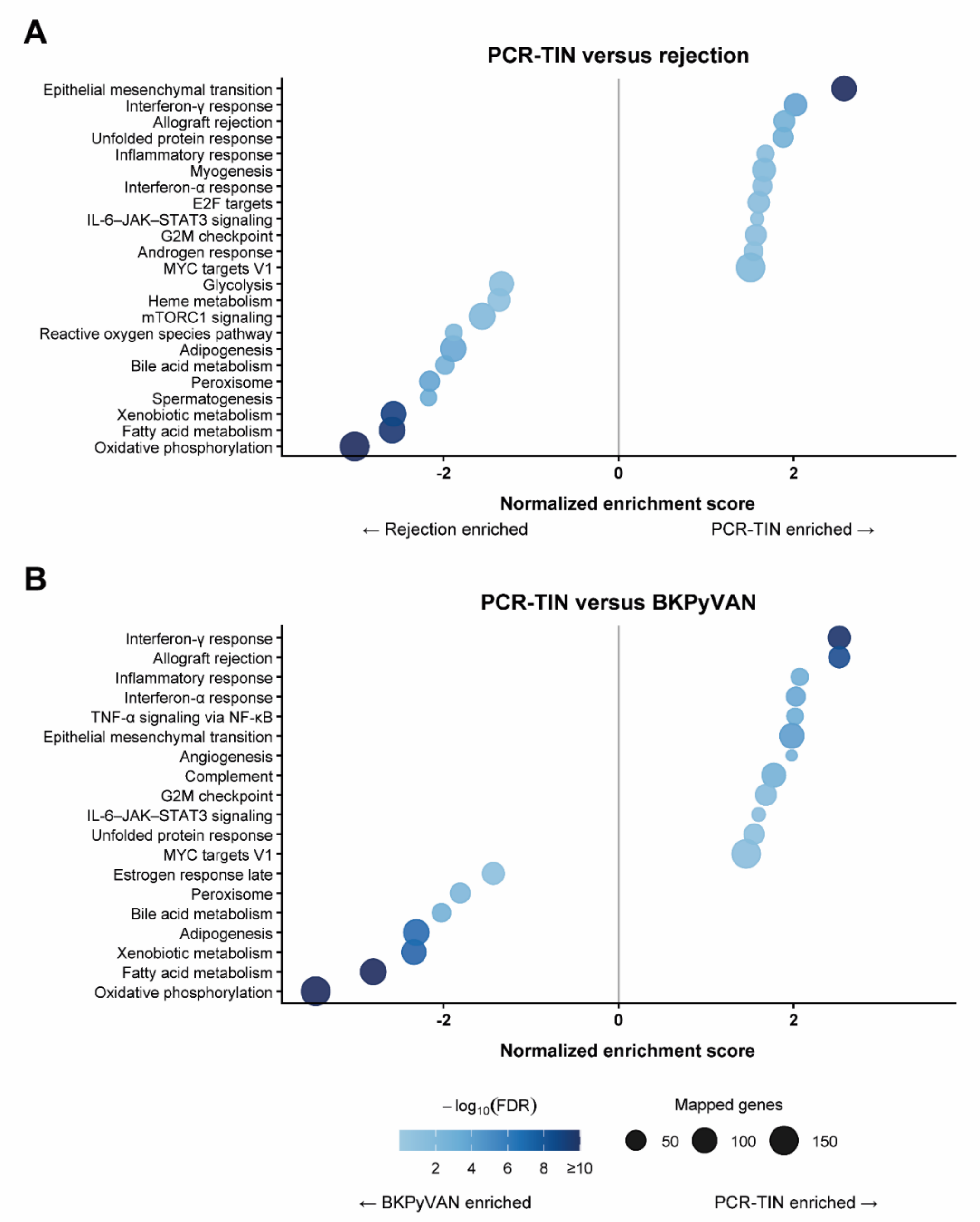


**Supplementary Figure S2. Hallmark Gene Set Enrichment in PCR-TIN Compared With Rejection and BKPyVAN.** Preranked gene set enrichment analysis of MSigDB Hallmark pathways **A.** PCR-TIN versus rejection. **B.** PCR-TIN versus BKPyVAN. Proteins were ranked using moderated t statistics from the corresponding limma differential-abundance analyses. Only pathways with a Benjamini–Hochberg FDR < 0.05 are shown. Positive normalized enrichment scores indicate enrichment in PCR-TIN, whereas negative scores indicate enrichment in rejection or BKPyVAN, respectively. Point size represents the number of mapped genes in each pathway, and color represents $-\log_{10}$(FDR), capped at 10 for visualization. The vertical line indicates a normalized enrichment score of zero.

**Abbreviations**: BKPyVAN, BK polyomavirus–associated nephropathy; FDR, false discovery rate; MSigDB, Molecular Signatures Database; PCR-TIN, plasma cell–rich virus-like tubulointerstitial nephritis.
